# Effects of Diet on the Microbiome and Serum Metabolome of South Asian Infants at 1 Year

**DOI:** 10.1101/2021.09.28.21264268

**Authors:** Colin Y. Bruce, Meera Shanmuganathan, Sandi M. Azab, Philip Britz-McKibbin, Sonia S. Anand, Russell J. de Souza, Jennifer C. Stearns

## Abstract

Diet is known to affect the gut microbiome and metabolome composition in adults, but this has not been fully explored in infants. Dietary patterns from 1 year-old infants (n=182) from the South Asian Birth Cohort (START) study were compared to gut microbiome alpha and beta diversity and to taxa abundance differences. Diet – serum metabolite associations were identified using multivariate analysis (partial least squares-discriminant analysis, PLS-DA) and univariate analysis (T-Test). Dietary biomarkers identified from START were also examined in a separate cohort of white Caucasian infants ([CHILD Cohort Study](https://childstudy.ca/), n=82). Lastly, the association of diet with gut microbiome and serum biomarkers, considering maternal, perinatal and infant characteristics was investigated using multivariate forward stepwise regression. A dietary pattern characterized by breastfeeding, supplemented by formula and dairy was the strongest predictor of the gut microbiome that also differentiated the serum metabolome of infants. The formula and dairy dietary pattern was associated with a panel of circulating metabolites in both cohorts, including: *S*-methylcysteine, branched-chain/aromatic amino acids, lysine, dimethylglycine, and methionine. Breastfeeding status, the prominent feature of the dietary pattern, was also associated with a sub-set of serum metabolites in both cohorts. In START, this diet pattern was associated with the metabolites tryptophan betaine, 2-hydroxybutyric acid, tyrosine, phenylalanine, and trimethyl-*N*-oxide. In the CHILD Cohort Study(CHILD), breastfeeding status was associated with the metabolites aminooctanoic acid, 3-hydroxybutyric acid, and methyl-proline. The results of our study suggest that breastfeeding has the largest effect on the composition of the gut microbiome and the serum metabolome at 1 year, even when solid food diet and other covariates are considered.

## 1. Introduction

Early life risk factors may contribute to chronic diseases of adulthood [1–3]. Several early life factors have been identified as predictors of future health outcomes. For example, rapid weight gain during early life increases risk of adult cardiovascular disease [3,4]. Infant vegetarianism or plant-based diets may promote immune system regulation through induction of T-regulatory cells and decrease risk of auto-immune disorders [5]. A longer duration of breastfeeding during infancy has been linked to decreasing incidence of gastrointestinal infections, asthma, and obesity [6,7]. Such diverse exposures may exert their effects through a common pathway—namely, the modulation of the gut microbiome [8]. The gut microbiome undergoes significant compositional changes during infancy and is emerging as an important predictor of long-term health [5,9] and chronic disease risk. Over the first year of life significant changes in diversity, richness, and relative abundances of gut microbes are observed as the microbiome matures and approaches an adult-like composition over the first 3-5 years of life [10–13]. A driver of gut microbial maturation is the infant diet, which also changes in composition from early life milk-based diets, towards adult-like solid food based diets [14]. Breastfeeding during early life supplies common milk substrates such as lactose and human milk oligosaccharides (HMOs) to the infant gut [10,14,15]. This substrate availability, especially HMOs, enriches for microbes characteristic of the early infant gut microbiome such as members of the genera *Bifidobacterium, Lactobacillus* and *Bacteroides*. These decrease in abundance at weaning [10,12,16]. The gut microbiomes of infants who are predominantly formula-fed exhibit a distinct microbiome composition from those who are breastfed, show lower abundances of *Bifidobacterium* compared to breastfed infants of the same age [15], and mature differently than the gut microbiomes of breastfed-dominant infants [14,17]. While the effects of breastfeeding and formula feeding on the infant gut microbiota have been well investigated, less is known about how the timing and nature of solid foods introduced influences this difference. The transition towards a solid food diet has been shown to increase the complexity of the gut microbiome, however, predominant feeding type may influence these processes [10,17].

Another potential mechanism for breastfeeding and early-life diet to influence health outcomes is via bioactive molecules mediating complex host-microbiome interactions. These metabolites may originate directly from specific foods in the diet [18], comprising both nutrient and non-nutrient food components where they can be classified as either *exposure biomarkers*, in which the food components exert some physiological effect on the host, or as *outcome biomarkers* following host and/or gut microbiota biotransformations [19]. A growing field of nutritional metabolomics investigates the relationship between diet and serum or urinary metabolites that may serve as objective *biomarkers of food intake* (BFIs) [20–23]. Alternatively, metabolites may be microbially-produced and secreted into the gut environment, such as trimethylamine-*N*-oxide (TMAO), which is derived from intake of fish [24] and/or synthesized via microbial metabolism of dietary carnitine or phosphatidylcholine [25]. Common substrates for microbial metabolism from dietary fibres also include short-chain fatty acids (SCFAs), notably acetate, butyrate, and propionate [26,27].

In this study, we investigated the association of host diet with the gut microbiome, diet with the serum metabolome, and the gut microbiome with the serum metabolome during the first year of life in a cohort of South Asian infants residing in the Greater Toronto Area (Ontario), which was also compared with a subset of white Caucasian infants from a pan-Canadian cohort.

## 2. Materials and Methods

### Study Design/participants

The South Asian Birth Cohort (START-Canada) enrolled 1012 South Asian mother–child pairs from the Peel Region of Ontario to investigate the influence of diverse environmental exposures and genetics on early life adiposity, growth trajectory, and cardiometabolic risk factors [28]. The infant was considered to be of South Asian ethnicity if both parents and grandparents’ ancestral origin was identified as from India, Pakistan, Sri Lanka, or Bangladesh. Participant data, including dietary data and microbiome data, for 182 samples were collected [8]. Serum metabolite data for 72 participants were also collected from START. For comparison of results across ethnicities, and for replication of the serum metabolite analysis, participant data (diet, microbiome, serum metabolite) for 82 participants that reported white Caucasian as their ethnicity was collected from CHILD [29]. CHILD participants included in this study were recruited across multiple centres in Canada (Toronto (n=21), Winnipeg (n=1), Edmonton (n=30), Vancouver (n=29)).

### Dietary data collection

Maternal and infant diet data were collected using validated semi-quantitative food frequency questionnaires (FFQs) [30–33]. The maternal semi-quantitative FFQ was developed and validated for use among South Asians living in Canada. On the FFQ, participants report how often, on average, they consumed selected foods in the previous year. Maternal diet data was collected during the second trimester of pregnancy. From the FFQ, we created 36 food groups as described previously [20,34], which we used to develop a diet quality score (DQS), calculated as the sum of daily servings of “healthy” foods (fermented dairy, fish and seafood, leafy green vegetables, cruciferous vegetables, legumes, fruits, nuts, and whole grains) less the sum of daily servings of “unhealthy foods” (processed meats, refined grains, French fries, snacks, sweets, and sweet drinks. These foods were chosen because they have been widely used to characterize healthy dietary patterns (i.e., prudent diet) that reduce chronic disease risk [35–42]. Our DQS correlates well with a modified version of the AHEI previously derived in these cohorts [20,34,43]. The infant semi-quantitative FFQ built upon existing instruments used in 3 studies: the Avon Longitudinal Study of Parents and Children (ALSPAC) [44], the Born in Bradford (BiB) study [45], and the Study of Health Assessment and Risk Evaluation (SHARE) [30]. These tools were developed for use in children, South Asian children, and South Asians living in Canada, respectively. Starting with items listed on any of the 3 instruments, dietitians and study personnel of South Asian origin refined the list to add missing foods, remove redundant foods, and produce a booklet that queried the infant’s frequency of consumption of 80 individual food items over the preceding 4 weeks. Respondents were asked to quantify the number of “servings/day”, “servings/week”, or “servings/month” as appropriate. Reference portion sizes were provided for each item, guided by the serving sizes in the ALSPAC instrument. The 80 food items were then sorted into common food groups resulting in 12 food groups: Refined Grains, Whole Grains, Processed Meats, Unprocessed Meats, Fish, Vegetables, Beans and legumes/nuts/plant proteins, Sweets, Fruits, Dairy, Eggs, and Other.

The infant diet questionnaire used for the CHILD cohort was similar to the one used with START. The CHILD infant diet instrument used a 13-item questionnaire asking about consumption of groups of foods that had been introduced in the first year of life. The 13 food items listed on the questionnaire were: Prepared baby food, Baby cereals, Commercial cereals, Grains, Dairy products, Vegetables, Fruits, Meats, Egg, Fish, Shellfish, Nuts, Peanuts/peanut butter.

### Health data collection

Health and other participant characteristic data included: maternal vegetarian status, weight gain over the first year of life, age in weeks at the 1 year sample collection, breastfeeding status at 1 year, current formula feeding status at 1 year, and offspring age in weeks when solid foods were introduced [1,34].

### Gut microbiome stool analysis

Sequence reads for the v3 region of the 16S rRNA genes from 182 infant stool samples from our previous study [8] were reprocessed for this study. Adapter, primer and barcode sequences were trimmed from sequencing reads with cutadapt [46], then amplicon sequence variants (ASV) were inferred from the sequenced data using DADA2 [47]. Taxonomy was assigned in DADA2 that uses the RDP naive Bayesian classifier method using the SILVA 16S rRNA gene reference file [48] (release version 132). Rarefied species richness [49] ACE, Chao1 and the inverse Simpson [50] and Shannon diversity were calculated from ASV count data with the vegan [51] package in R. Between-community diversity analyses were based on the Aitchison distance [52] calculated using the Euclidean distance of centre-log transformed ASV level count data. A PCA of Aitchison distance was calculated with x to generate an eigen value for each sample from PC1 and PC2. Bacteroidetes:Firmicutes ratios were calculated for each sample by agglomerating taxa at the phylum level for each sample and dividing the total Bacteroidetes read counts for each sample by the total Firmicutes read counts. The same process was done for calculating the Firmicutes:Actinobacteria ratio.

### Serum sample pre-treatment and instrumental analysis for serum metabolomics

Frozen serum samples (START (n=72), CHILD (n=82)) were thawed on ice and an aliquot of 50 µL was transferred into a 0.5 mL centrifuge tubes for sample dilution (a 4-fold) with internal/recovery standards followed by ultrafiltration (3 kDa) to remove protein (i.e., serum filtrate) as described previously [53]. In addition, a pooled quality control (QC) was prepared by combining an equal aliquot of serum samples from START maternal cohort (n=300) that was used to assess technical precision (This was the first cohort to be analyzed in the lab). All serum filtrate and QC samples were stored frozen at -80°C prior to metabolomic analysis by multisegment injection-capillary electrophoresis-mass spectrometry (MSI-CE-MS). A standardized protocol was used to measure polar/ionic serum metabolites under positive and negative ion mode detection with full-scan data acquisition [53]. This technique takes advantage of a serial sample injection format of seven (or more) samples within a single run [54] using an Agilent 6230 time-of-flight mass spectrometer (TOF-MS) with a coaxial sheath liquid Jetstream electrospray ion source coupled to an Agilent G7100A capillary electrophoresis (CE) unit.

Following data acquisition, all raw MSI-CE-MS data (.d format) were processed using MassHunter Workstation Qualitative Analysis software (version B.06.00, Agilent Technologies, 2012). Both a targeted (i.e., known metabolites confirmed with reference standards) and a nontargeted (i.e., unknown ions authenticated from molecular features and putatively identified by MS/MS) workflows were used to characterize the infant serum metabolome [53], where all ions were annotated based on their accurate mass-to-charge ratio (m/z), relative migration time (RMT) and mode of detection (p:positive, n:negative). Peak areas and migration times for all serum metabolites and internal/recovery standards were transferred to an Excel worksheet (Microsoft Office) and relative peak areas (RPA) for each unique molecular feature were exported as a .csv file. Serum metabolites frequently detected in > 75% of all samples analyzed with adequate technical precision (CV < 30% for repeat QC samples analyzed) were included in the data matrix for further statistical analysis. Missing value inputs were replaced by a value equal to one-half the lower limit of detection. Serum metabolite concentrations (□M) were reported for all known/confirmed serum metabolites, whereas relative peak area (RPA) were used for tentatively identified or unknown metabolites lacking available standards.

### Statistical analysis

Linear regression models were fitted to test the association of diet patterns and participant characteristics with gut microbiome alpha diversity. Univariable tests of the association of diet pattern, and participant characteristics with gut microbiome beta diversity were done with PERMANOVA with 99,999 permutations using the vegan package in R. Multivariable models to assess the effect of diet patterns and participant characteristics on gut microbiome beta diversity were fitted with the envfit function in the vegan package. Bacterial ASV enrichment based on the formula and dairy dietary pattern were measured with the DESeq2 package in R [55].

To assess the relationship between diet and the serum metabolome, diet patterns were compared to serum metabolite concentrations using the Metaboanalyst [56] web platform. Serum metabolite data were *log*-transformed and auto-scaled prior to supervised multivariate data analysis using partial least squares-discriminant analysis (PLS-DA). A variable importance in projection (VIP) plot was used to rank order serum metabolites of significance to the PLS-DA model. Metabolites that had the greatest contribution to the differences between groups in the PLS-DA model were carried forward for further investigation. For START samples univariate testing (Tukey’s T-test, FDR adjusted for multiple hypothesis testing) was used to confirm the significance of serum metabolites with diet patterns and an adjusted *p* < 0.05 was considered significant. Univariate metabolite analysis was adjusted for covariates listed in Table S4. A similar analysis was done to compare serum metabolites associated with breastfeeding status in START participants at 1-year collection time. To confirm the associations of serum metabolites with breastfeeding status the same analysis was repeated using samples from CHILD. The association of diet and participant characteristics with the concentration of serum metabolites of interest was modeled with univariate and multivariate models that included a combined dataset from START and CHILD. Variables significant at *p* < 0.1 in univariate testing were carried into a forward stepwise regression model.

### Self-reported dietary intake

We took a targeted approach to assessing the association between infant food intake and circulating metabolite concentrations in non-fasting serum. We started with a list of previously identified robust dietary biomarkers of specific foods in adults [20,24] within this infant population. These included proline betaine (citrus fruits), methylhistidine (red meat, chicken, eggs), hippuric acid (vegetable, fruit), TMAO (seafood, meat, red meat, eggs), and tryptophan betaine (nuts, seeds, peanuts, legumes).

We explored several approaches to characterizing the diet in this analysis, as follows: 1) We created a diet diversity index (“dietary diversity 1”), which was a simple count of the number of different food groups (i.e. out of 36 for mothers; out of 12 for infants) that an individual reported consuming a non-zero amount of; 2) We created a diet richness index (“dietary diversity 2”) in which we simply counted the number of items (i.e. out of 152 for mothers; out of 80 for infants) that were consumed a non-zero amount of times); 3) We used principal components analysis (PCA) to derive dietary pattern scores for mothers and infants (as described previously [34]); 4) We used a PCA to derive 3 dietary patterns for infants, using the 12 food groups; these patterns were called animal foods, plant foods, and infant formula and dairy.

## 3. Results

### Description of the infant diet

The interquartile range for diet diversity 1 was 7 to 9; and for diet diversity 2, 2 to 3. Three distinct dietary patterns were retained from the unsupervised PCA model, 1: Animal foods (unprocessed meat, processed meat, fish, eggs; eigenvalue = 1.43), 2: Plant foods (whole grains, vegetables, beans/legumes, fruits, tea/water; eigenvalue = 1.16), 3: Formula/dairy (dairy, formula, with a negative loading on breastfeeding; eigenvalue = 0.92), which together explained 44.6% of the variance in diet.

### Diet and participant characteristics associated with infant gut microbiome composition

We investigated the association between infant variables and gut microbial alpha diversity from START participants and found that infant weight gain over the first year of life was negatively associated with Shannon diversity (−0.063, *p* = 0.009), however, no association was found between diet diversity or the diet patterns and microbial alpha diversity estimates. Comparison of the same set of diet variables and participant characteristics to beta diversity (Aitchison distance) identified that number of items eaten during the reporting period and age in months at 1 year collection time were associated with beta diversity in univariable PERMANOVA analysis (Table 1). The multivariable *envfit* model indicated that adherence to the formula and dairy dietary pattern and age were associated with beta diversity (Table 1). For *envfit* analysis the breastfeeding status was not included in the same model with adherence to the formula and dairy dietary pattern because both variables include breastfeeding as a component, and thus were highly collinear (r = -0.77). In fact, low adherence to the formula and dairy dietary pattern corresponded to current breastfeeding status, and high adherence corresponded to 2.22 servings of formula and 6.67 serving of dairy.

**Table 1.** Beta Diversity Scores Associated with Dietary Variables

| Variable | Univariable<br>R<sup>2</sup> (P-Value) | Multivariable<br>R<sup>2</sup> (P-value) |
| --- | --- | --- |
| Dietary Diversity | 0.006 (0.166) | 0.034 (0.754) |
| Number of Items Eaten | **0.008 (0.009)** | 0.026 (0.519) |
| Adherence to Animal Foods DP | 0.006 (0.243) | 0.003(0.930) |
| Adherence to Plant Foods DP | 0.006 (0.229) | 0.013(0.350) |
| Adherence to Formula and Dairy DP | **0.007 (0.078)** | **0.109 (<0.001)** |
| Breastfeeding status | **0.008 (0.004)** |  |
| Mother Vegetarian Status in Pregnancy | 0.006 (0.182) | 0.017(0.189) |
| weight gain in the first year | 0.007 (0.062) | 0.007(0.536) |
| Age in months at collection | **0.008 (0.014)** | **0.046 (0.015)** |
| Age when solid foods introduced | 0.006 (0.381) | 0.031(0.197) |

DP = dietary pattern

We next looked at the the enrichment of gut microbial taxa based on adherence to the formula and dairy dietary pattern and found that many ASV from *Lactobacillus, Bifidobacterium, Veillonella* and *Megasphaera* were less abundant in infants with a high compared with a low adherence to the formula and dairy dietary pattern (Table S1). We investigated whether these associations were driven by solid food consumption differences between breastfed and not-breastfed infants but we did not find significant differences in the amounts of commonly consumed foods such as legumes (difference = 0.07 servings/d, p-value = 0.34), and eggs (difference = -0.12 servings/d, p-value = 0.18). We also found an enrichment in high adherence to the formula and dairy dietary pattern of ASV from Enterobacteriaceae, *Enterobacter, Lactococcus, Blautia, Erysipelatoclostridium* (Table S1).

### Associations of the Infant Diet with the Infant Serum Metabolome

To observe how diet at 1 year may be influencing the serum metabolome, a supervised PLS-DA model was applied to compare differences in the serum metabolic phenotype of infants as a function of their dietary pattern adherence score. We included serum samples from infants with low adherence (n = 26), medium adherence (n= 17), or high adherence (n= 29) to the formula and dairy dietary pattern, and observed almost complete overlap of profiles from medium and high adherence groups, but a distinct metabolome profile for low adherence infants (Figure 1A).

**Figure 1.**
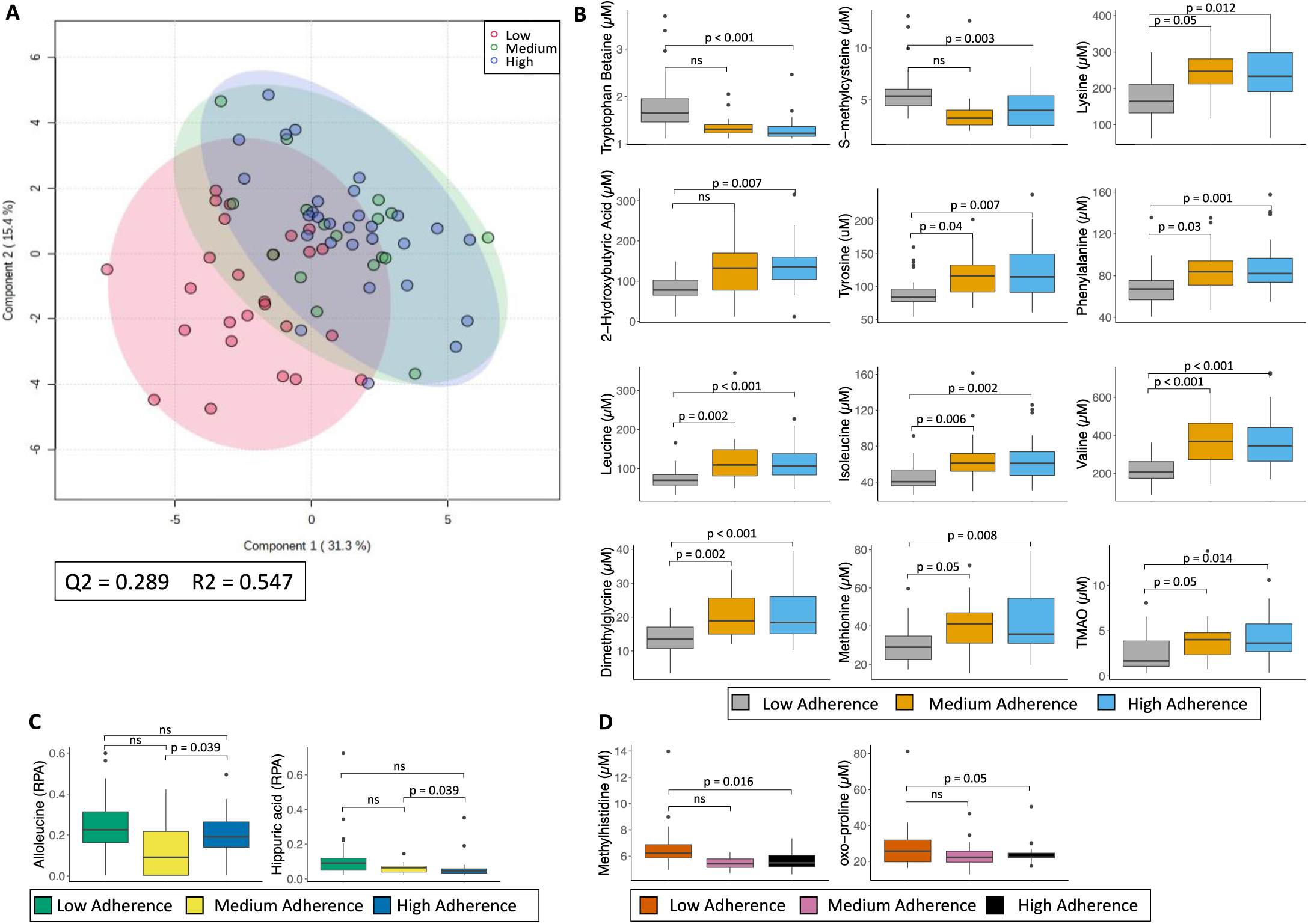
Metabolite concentrations related to dietary pattern adherence in the START cohort. **A)** PLS-DA of samples related to formula and dairy dietary pattern adherence with 95% confidence interval. Cross-validation was performed using leave-out-one-at-a-time cross-validation, and Q2 was maximized with 2 principal components. Serum metabolites related to **B)** formula and dairy dietary pattern adherence, **C)** animal foods dietary pattern adherence and **D)** plant foods dietary pattern adherence. Boxplots are coloured by adherence to specific diet pattern at collection time (box = 1st and 3rd quartiles with median, wiskers = 1.5x IQR, dots = outliers). P-values are from Tukey’s T-tests, FDR adjusted. For the animal and plant dietary patterns, samples from infant who were breastfeeding status were removed prior to analysis. RPA: relative peak area

Separation of infant serum metabolite profiles was primarily due to dimethylglycine (DMG), valine, and tryptophan betaine (VIP > 1.5; Figure S1). Univariate (Tukey’s T-test, FDR adj. *p* < 0.05) tests confirmed the associations of these metabolites of interest with adherence to the formula and dairy diet pattern, as well as phenylalanine, *S*-methylcysteine, leucine, tyrosine, 2-hydroxybutyric acid, and lysine (Table S2). Low adherence to the formula and dairy dietary pattern at 1 year was associated with higher concentrations of tryptophan betaine and *S*-methylcysteine, whereas medium and high adherence to the formula and dairy dietary pattern was associated with higher lysine, 2-hydroxybutyric acid, dimethylglycine, methionine, TMAO, tyrosine and phenylalanine (aromatic amino acids), leucine and valine (branched-chain amino acids; BCAA) (Figure 1B). Tryptophan betaine is a serum biomarker of legume [20,57] and nut intake [24,58], and legume consumption was high in the START cohort [34]. We found no association between circulating tryptophan betaine in infant serum and either maternal or infant legume and nut intake (data not shown).

We next excluded infants who were breastfeeding at the time of the sample collection from the analysis and compared the serum metabolomic profile based on the adherence to the plant foods dietary pattern and the animal foods dietary pattern. We used univariate (Tukeys’ T-test, FDR adj. *p* < 0.05) tests on any metabolite with a VIP score ≥ 1.5 and found that high adherence to the plant foods dietary pattern was associated with higher alloleucine and hippuric acid than medium adherence, but was not significantly different from low adherence (Figure 1C). Adherence to the animal foods dietary pattern was associated with a lower concentration of methylhistidine and oxo-proline (Figure 1D). Since low adherence to the formula and dairy dietary pattern in START was associated with a greater likelihood of breastfeeding, we looked at the association of serum metabolites with breastfeeding in START. The same approach was used comparing START participants that were breastfed at collection time (n = 31) to non-breastfed at collection time (n = 41) that generated a PLS-DA model similar to the formula and dairy diet pattern (Figure S2). The breastfed group overlaps closely with the low adherence to the formula and dairy diet pattern group, and the non-breastfed group with the medium and high adherence groups. As expected, the same serum metabolites were determined to differentiate infant metabolite profiles (tryptophan betaine, dimethyl-glycine, valine, VIP > 1.5; Figure S2) as a function of breastfeeding adherence patterns. The two groups shared the same top 11 metabolites as ranked by VIP scores (VIP > 1.1), with only TMAO not as strongly associated with breastfeeding status as compared to the formula and dairy diet pattern.

To determine whether serum metabolic phenotypes associated with breastfeeding status that had been observed were specific to START, the same analysis was repeated in an independent cohort of white Caucasian 1-year old infants from CHILD (*n*=117) having otherwise similar characteristics (Table S3) as the infants in START. The results from START were then replicated in CHILD for 7 of 11 serum metabolites (Tukey’s T-test, adj. *p* < 0.05). The concentration of *S-*methylcysteine was higher in breastfeeding infants than non-breastfeeding infants, whereas leucine, isoleucine, valine, lysine, dimethylglycine, and circulating methionine was lower (Figure 2A). We then investigated additional association of serum metabolites with breastfeeding status in CHILD alone, in order to identify metabolites unique to this population, which revealed that serum 3-hydroxybutyric acid was higher in breastfed infants, whereas aminooctanoic acid and *N*-methylproline were lower in white Caucasian infants not breastfeeding (Figure 2B).

**Figure 2.**
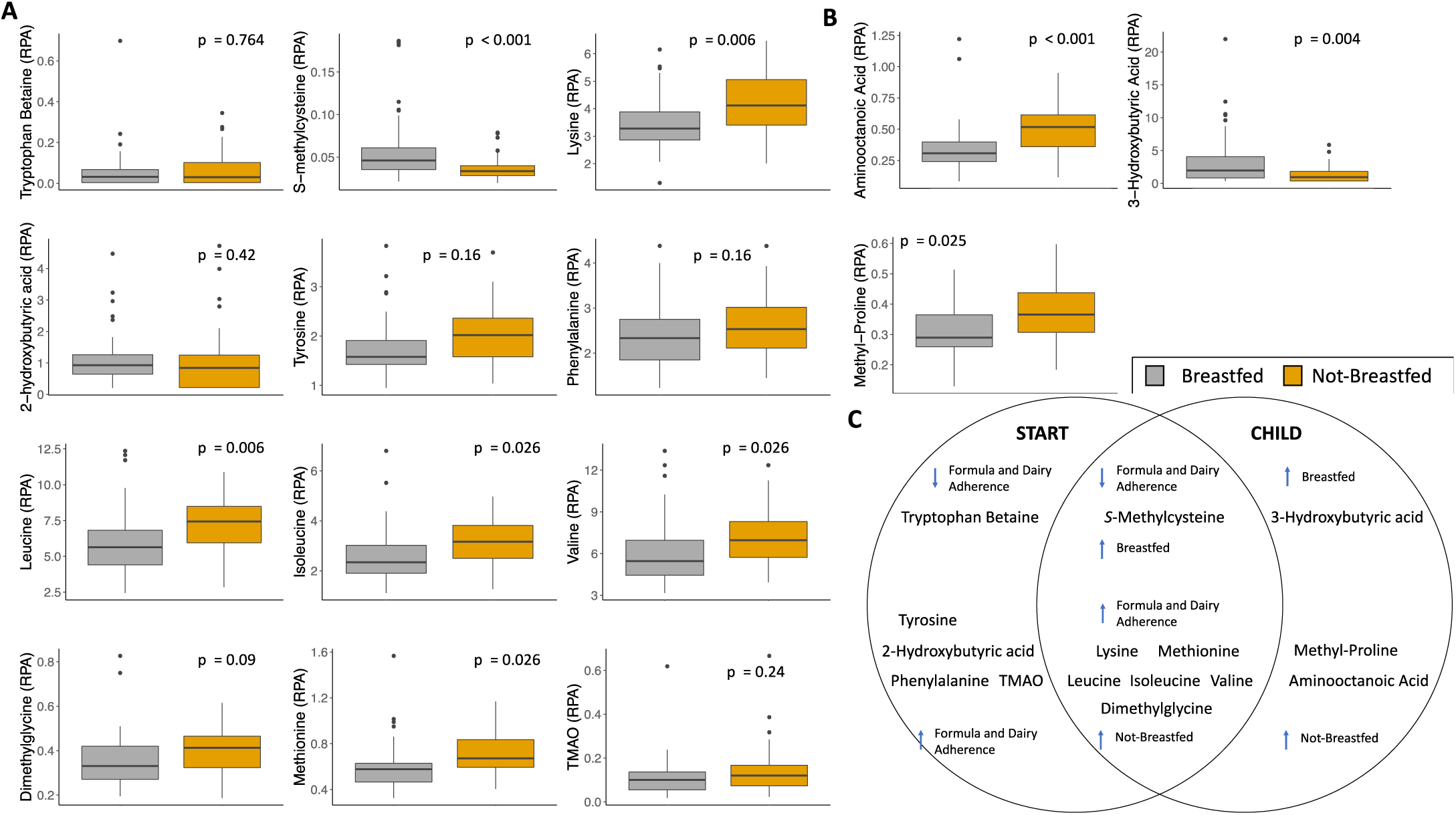
Metabolite concentrations within the CHILD cohort. **A)** Serum metabolites investigated since they were associated with the formula and dairy dietary pattern in START. **B)** Serum metabolites uniquely associated with breastfeeding status in CHILD participants. Boxplots of serum metabolite concentrations are coloured by participant breastfeeding status at sample collection time (box = 1st and 3rd quartiles with median, wiskers = 1.5x IQR, dots = outliers). P-values are from Tukey’s T-tests, FDR adjusted. **C)** Venn diagram of serum metabolites associated with diet in each study cohort. Arrows indicate the directionality of the association of metabolite concentrations with diet.

### Triangulating the associations between infant diet, gut microbiome and serum metabolome

Having found associations between the infant diet and the gut microbiome and associations between the infant diet and the serum metabolome, we then explored the contribution of diet and the gut microbiome to the concentrarion of specific serum metabolites while accounting for maternal, perinatal and infant characteristics. We tested whether serum concentration levels of metabolites significantly associated with diet in both cohorts: *S*-methylcysteine, leucine, isoleucine, valine, lysine, methionine, and dimethylglycine, were explained by feeding-related factors (breastfeeding status at 1 year, time since weaning, formula feeding status at 1 year, age of solid food introduction and cows milk consumption), the gut microbiome (PC1, PC2, Bacteroidetes/Firmicutes ratio and Firmicutes/Actinobacteria ratio), maternal characteristics (years in Canada), perinatal variables (GDM, delivery mode, maternal antibiotic and vegetarian status during pregnancy), or infant characteristics (birth weight, ethnicity, sex, weight gain in the first year and age) using forward stepwise regression (Table S4). Breastfeeding status was associated with all metabolites investigated following forward stepwise regression. *S*-methylcysteine, valine, leucine, and isoleucine were most strongly associated with breastfeeding status. *S*-methylcysteine concentration was only associated with breastfeeding status, leucine and valine concentrations were associated with breastfeeding status and microbiome PC2, and isoleucine was associated with breastfeeding status and maternal antibiotic use during pregnancy (Table S4). Lysine, methionine, and DMG were most strongly associated with microbiome PC2, followed by breastfeeding status. Lysine concentrations were also associated with maternal vegetarian status, and DMG was associated with formula feeding status at 1 year collection time.

## 4. Discussion

In this study, we have explored the complex interactions of infant diet in relation to infant serum metabolome and the infant gut microbiome in order to better understand the role of gut microbial communities in infant nutrition and metabolism within a Canadian cohort infants of South Asian and European descent. The results of this study indicate that gut microbiome diversity of 1-year old infants is affected by breastfeeding status at 1 year, but not by solid food consumption. The infant serum metabolome was influenced by host diet, with distinct serum metabolites identified with relation to consumption of animal foods, plant foods, and most strongly with infant formula and dairy foods.

The formula and dairy dietary pattern was associated with a panel of circulating metabolites in both cohorts, including: *S*-methylcysteine, branched-chain/aromatic amino acids, lysine, dimethylglycine, and methionine. The results of our study suggest that breastfeeding has the largest effect on the composition of the gut microbiome and the serum metabolome at 1 year, even when solid food diet and other covariates are considered.

Our multivariate analysis results find that breastfeeding status and infant formula and dairy consumption are the drivers of gut microbiome composition, regardless of the types of solid foods consumed in the first year of life. These results are supported by specific taxa enrichment in response to dietary variables in which microbes associated with breastfeeding, largely belonging to the genera *Bifidobacterium, Lactobacillus* and *Veillonella* were consistently less abundant with increasing formula and dairy diet pattern adherence. The lower abundance of *Bifidobacterium* and *Lactobacillus* in response to cessation of breastfeeding is likely due to altered substrate availability, and is consistent with previous studies that showed similar decreases in both genera [10]. Increasing formula and dairy consumption was associated with a higher abdundace of the genus *Blautia*, which are characteristic of late infancy and adult populations [10]. This increase is likely in response to the transition away from breastfeeding because *Blautia* are unable to metabolize HMOs [59][60].

Infant age was also a predictor of microbial β diversity. Age has been previously considered to be an important factor in the maturation of the gut microbiome [8,10,17]. Previously, infant formula feeding has shown to produce a more adult-like gut microbiome than breastfeeding [10]. Weaning from breastmilk and the transition to solid foods has also been suggested to be a driver of gut microbial change [10].

We identified tryptophan betaine and *S*-methylcysteine as dietary biomarkers associated with breastfeeding status among infants, with tryptophan betaine uniquely associated with breastfeeding in South Asian participants. Previous studies have detected tryptophan betaine in breastmilk samples [61], suggesting its transfer into the infant gut through breastfeeding. Future work may include testing samples of breastmilk for the presence of these metabolites. Tryptophan betaine, also known as hypaphorine, is an indole alkaloid that has been previously measured in nuts and legumes (e.g., chickpeas and lentils) [20,24,57]. A small study in breastfeeding mothers (n =24) reported its presence in breast milk and also an association with peanut consumption [61]. Dietary challenge in lactating women with hypaphorine-rich foods (restricted intake for 3 days before the intervention) demonstrated transfer of hypaphorine into milk with hypaphorine appearance peaking 5-18 h after consumption. Prolonged disappearance of hypaphorine from the milk was indicative of slow excretion or metabolism (over 2-3 days) [61]. This suggests that there may be an ideal window to capture BFIs in the infants serum. We previously reported a strong association between nut and legume consumption and measured serum tryptophan betaine concentrations in pregnant women from START (n=300) [20]. However, in the present analyses, we did not detect an association of maternal nut and legume intake with infant serum tryptophan betaine (0.007, p = 0.64),. The biomarkers presented here reflect cultural dietary practices, such as vegetarianism and habitual intake of legume-rich foods among the South Asians in our study. Infant nursing practices may also play a part in these differences, as START infants were more commonly formula fed or mixed-fed at collection time compared to white Caucasian infants from CHILD [8]. Identifying dietary biomarkers may allow a more tailored approach for investigating infant nutrition and the potential sources of nutrients, as they may originate from maternal or infant diets.

*S*-methylcysteine was shown to be strongly associated with breastfeeding status among both cohorts, independently of ethnicity, suggesting a potential biomarker of breastfeeding. *S*-methylcysteine has previously been reported to increase in host serum following consumption of navy beans, garlic, and onions [62–64]. *S*-methylcysteine may be related to host health benefits including reducing risk of cancer, and potentially anti-inflammatory activity [62,65]. Previous reports have not linked *S*-methylcysteine concentrations to breastfeeding. It is possible that this metabolite behaves similarly to tryptophan betaine, introduced through consumption of beans and passed to the infant through breastmilk. Correlational analysis between *S*-methylcysteine and other serum metabolites identified moderate correlations with glycine (r = 0.44), glutamine (r = 0.41), tryptophan betaine (0.40), suggesting these as potential co-occuring metabolites, as well as weak correlations with serine (r = 0.34) and choline (r = 0.30). Glycine has been reported to be common in vegetarian and vegan diets, and is commonly consumed in soy products [66,67]. Glutamine is an amino acid commonly found in meat products, but may also be found in plant foods including soy and peanuts [68]. Tryptophan betaine, as we have previously described, is commonly found in nuts and legumes [20,24,57]. Serine is a precursor for *S*-methylcysteine in beans, and acts as a precursor for glycine production in humans [69,70]. Choline is commonly found in animal products, but is also found in beans and soy products [62]. The sources of metabolites that are postively correlated with *S*-methylcysteine concentration suggest the source of this metabolite to be from beans and bean product consumption. The relationship of *S*-methylcysteine with breastfeeding is still unclear, but may represent a mode of transfer to the infant.

Serum metabolites that were associated with adherence to the plant and animal foods diets have been previously identified in foods related to those patterns. Hippuric acid, identified to be in higher concentration in participants who consumed a high plant foods adherent diet, has previously been shown to be prevalent in a variety of plant foods [20,22,71]. Similarly, methylhistidine is found in a number of animal products and is related to protein turn-over [20,21,24,72–74].

The serum metabolites that were inversely associated with breastfeeding status in the START cohort included dimethylglycine, valine, phenylalanine, leucine, isoleucine tyrosine, lysine, methionine, TMAO, and 2-hydroxybutyric acid. The same associations were replicated in CHILD for the metabolites valine, leucine, isoleucine, lysine, and methionine.

Two metabolites were inversely associated with breastfeeding status and were unique to the CHILD cohort, aminooctanoic acid and 3-hydroxybutyric acid. 3-hydroxybutyric acid (3-hba) is a ketone body, commonly associated with exercise and periods of fasting [75]. 3-hba may be a marker of type II diabetes, when it is significantly higher in plasma compared to control groups [76]. It may also have a role in repressing immune activated inflammation related to obesity and aging [77].

The identification of the BCAAs leucine, isoleucine, and valine, may allow them to serve as biomarkers of formula consumption in infants. The prevalence of BCAAs and TMAO in the serum of formula fed infants may be due to higher protein and choline concentration in infant formulas compared to breastmilk [78–80], especially breastmilk produced when the infant is 1 year of age.

The infant gut microbiome, particularly the presence of *Bifidobacterium* during early life, has been associated with development of the immune system, prevention of allergy, and the production of nutrients available to the host [10,81]. The identification of objective dietary biomarkers for breastfeeding, formula feeding and the solid food diets will enhance our understanding of the contribution of the infant diet to metabolism and development. Future studies should investigate how dietary biomarkers are introduced into the infant serum, and whether they may be passed from mother to offspring during breastfeeding, as well as their effects on offspring health.

### Strengths and limitations

This study investigated a unique population of infants, the South Asian population, that has dietary characteristics distinct from white Caucasian populations, for instance the prevalence of vegetarianism and the higher rates of formula and mixed feeding. A strength of this study is the validation of diet-metabolite associations in a second cohort of white Caucasian infants with the identification of ethnic-specific metabolites. Importantly, we replicate the association of dietary biomarkers previously validated in the adult population in infants. Potential weaknesses of this study are the small sample sizes used for the metabolite analysis, as only 153 infants in this study had serum samples collected, which were collected under non-fasting conditions. In addition, only polar ionic metabolites were analyzed. Future work should be expanded to lipid classes of metabolites, and to include analysis of lipids from serum extracts as dietary biomarkers of full-fat dairy intake [82].

## 5. Conclusions

In this cohort of 72 South Asian infants aged 1 year, breastfeeding status has the largest effect of all dietary variables on gut microbiome composition. Infant formula consumption, dairy consumption, and other animal food consumption lead to a decrease in gut microbes characteristic of the healthy infant gut microbiome, primarily *Bifidobacterium*. Plant based diets may prevent the decrease in *Bifidobacterium* seen with other solid food diets. Serum tryptophan betaine was associated with breastfeeding status only in South Asian infants, wheras serum *S*-methylcysteine and branched-chain amino acids were associated with breastfeeding status and formula feeding status respectively, across two cohorts, and may serve as generalizable dietary biomarkers for infants.

## Data Availability

The datasets generated and/or analyzed during the current study are not publicly available, since the CHILD and START studies are bound by consent and cannot provide identifiable information to an outside group, but are available from the corresponding author on reasonable request.

## Author Contributions

Conceptualization, S.S.A and J.C.S; methodology, R.J.d.S, P.B-M, and J.C.S; formal analysis, C.Y.B; resources, P.B-M and J.C.S; data curation, M.S, P.B-M, and R.J.d.S.; writing—original draft preparation, C.Y.B and J.C.S; writing—review and editing, All; visualization, C.Y.B; supervision, SSA; project administration, S.S.A; funding acquisition, S.S.A. All authors have read and agreed to the published version of the manuscript.

## START study Team Acknowledgements

South Asian Birth Cohort (START) study data were collected as part of a bilateral program funded by the Indian Council of Medical Research/Canadian Institutes of Health Research (grant INC-109205) as well as additional funding from Heart and Stroke Foundation of Canada grant NA7283. Dr Sonia Anand and Dr. Milan Gupta were the founding co-PIs for START. Sonia Anand is supported by a Tier 1 Canada Research Chair in Ethnic Diversity and Cardiovascular Disease, and a Heart and Stroke Foundation/Michael G. DeGroote Chair in Populaiton Health Research at McMaster University.

## Acknowledgments

We thank the CHILD Cohort Study (CHILD) participant families for their dedication and commitment to advancing health research. CHILD was initially funded by CIHR and AllerGen NCE. Visit CHILD at childcohort.ca

