## Supplemental Table 1 for "Effects of Diet on the Microbiome and Serum Metabolome of South Asian Infants at 1 Year"

**Table S1.** DESeq2 results for adherence to the formula and dairy dietary pattern (high vs. low)

| | **Base Mean** | **Log2 Fold Change** | **Std Err.** | **p** | **FDR adj p** |
|---|---|---|---|---|---|
| ASV266 Enterobacteriaceae | 7.84 | 22.03 | 1.71 | 6.02E-38 | 9.80E-36 |
| ASV138 *Enterobacter* | 19.61 | 4.97 | 1.68 | 0.003 | 0.038 |
| ASV46 *Lactococcus* | 115.84 | 4.36 | 0.99 | 1.01E-05 | 2.74E-04 |
| ASV6 *Blautia* | 3856.83 | 1.81 | 0.53 | 7.01E-04 | 0.010 |
| ASV100 *Erysipelatoclostridium* | 82.74 | 1.77 | 0.50 | 3.93E-04 | 0.006 |
| ASV157 *Lactobacillus* | 64.77 | -8.84 | 1.29 | 7.94E-12 | 6.47E-10 |
| ASV184 *Lactobacillus* | 10.98 | -8.80 | 1.68 | 1.61E-07 | 6.55E-06 |
| ASV68 *Lactobacillus* | 99.37 | -7.43 | 1.92 | 1.04E-04 | 0.002 |
| ASV304 *Megasphaera* | 16.62 | -7.30 | 1.46 | 5.45E-07 | 1.78E-05 |
| ASV161 *Lactobacillus* | 18.40 | -6.64 | 1.75 | 1.45E-04 | 0.003 |
| ASV8 *Bifidobacterium* | 3005.68 | -4.09 | 1.12 | 2.46E-04 | 0.004 |
| ASV60 *Veillonella* | 163.39 | -3.56 | 0.90 | 7.99E-05 | 0.002 |
| ASV67 *Veillonella* | 195.07 | -2.50 | 0.87 | 0.004 | 0.047 |
| ASV11 *Bifidobacterium* | 4104.51 | -2.48 | 0.43 | 1.05E-08 | 5.70E-07 |
