## Supplemental Figure 2 for "Effects of Diet on the Microbiome and Serum Metabolome of South Asian Infants at 1 Year"

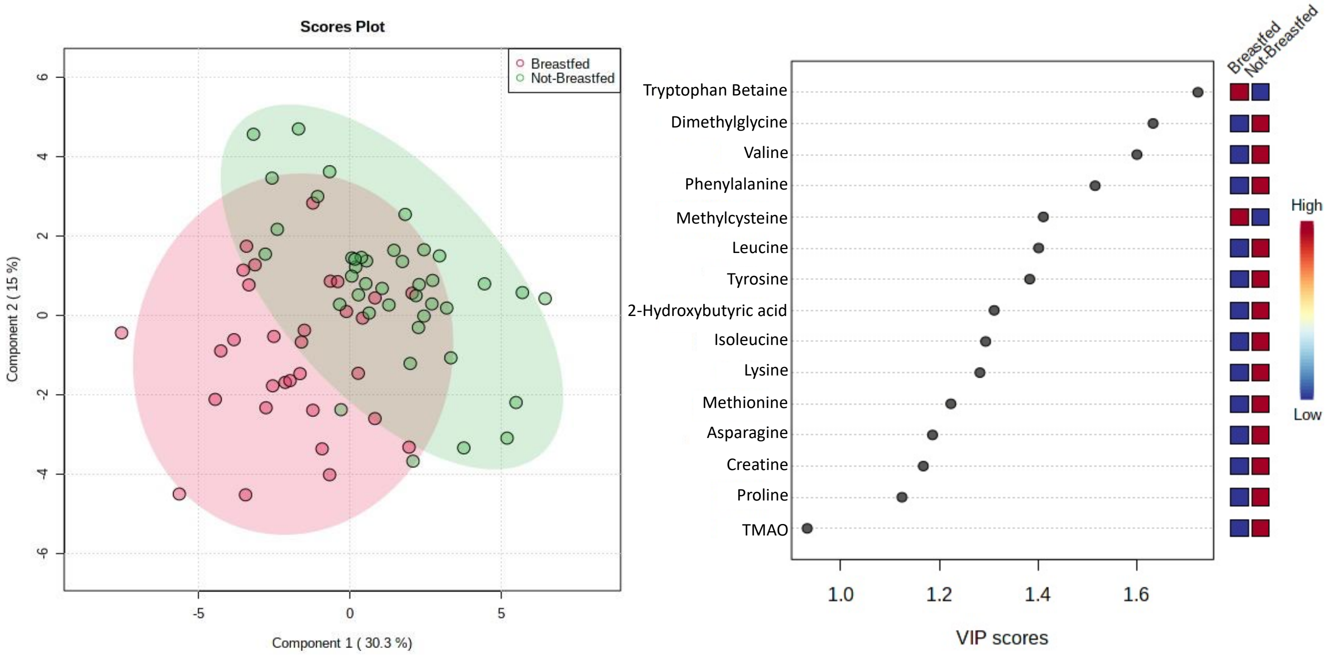


**Figure S2.** PLS-DA of breastfeeding status at 1 year collection time. Serum metabolite profiles for participants (n = 72) were generalized log-transformed and auto-scaled prior to PLS-DA production. Participant profiles were grouped based on breastfeeding status at 1 year collection time, and are coloured and grouped via 95% confidence interval based on this status. Metabolites responsible for the generation for the PLS-DA projection are listed on the right, with variable importance in projection (VIP) scores shown on the x-axis, as well as concentration of the metabolites with breastfeeding status.
