## Supplemental Figure 1 for "Effects of Diet on the Microbiome and Serum Metabolome of South Asian Infants at 1 Year"

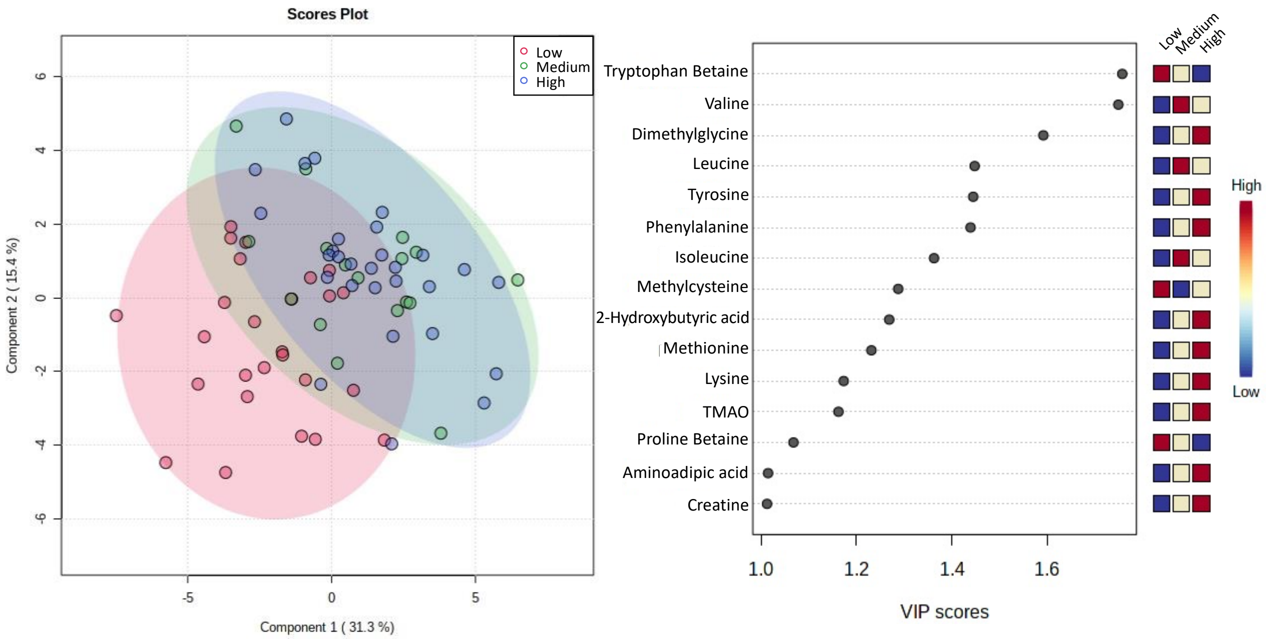


**Figure S1**. PLS-DA of adherence to the formula and dairy diet pattern. Serum metabolite profiles for participants (n = 72) were generalized log-transformed and auto-scaled prior to PLS-DA production. Participant profiles were grouped into 3 tertiles of adherence to the formula and dairy diet pattern and are coloured based on this adherence, as well as grouped via 95% confidence interval. Metabolites responsible for the generation for the PLS-DA projection are listed on the right, with variable importance in projection (VIP) scores shown on the x-axis, as well as concentration of the metabolites with each adherence group.
