## Supplemental Table 4 for "Effects of Diet on the Microbiome and Serum Metabolome of South Asian Infants at 1 Year"

**Table S4**. Forward stepwise regression of serum metabolites with diet, microbiome, maternal, perinatal and infant characteristics

| | | S-methylcysteine | | Leucine | | Isoleucine | | Valine | |
|---|---|---|---|---|---|---|---|---|---|
| | Variable | Univar. | FSW | Univar. | FSW | Univar. | FSW | Univar. | FSW |
| Infant diet | Breastfeeding Status at 1 year | **0.0184 (0.000)** | **0.0161 (0.001)** | **-2.099 (0.000)** | **-1.743 (<0.001)** | **-0.730 (0.000)** | **-0.627 (0.002)** | **-1.820 (0.000)** | **-1.603 (0.001)** |
| | Time since Weaning | **-0.0008 (0.097)** | | **0.221 (0.021)** | | **0.094 (0.018)** | | 0.130 (0.106) | |
| | Formula Feeding Status at 1 year | **-0.0087 (0.057)** | | -0.044 (0.931) | | 0.071 (0.740) | | 0.345 (0.445) | |
| | Age of Solid Food Introduction | 0.0029 (0.464) | | 0.358 (0.774) | | 0.438 (0.406) | | 0.242 (0.827) | |
| | Cow Milk Consumption | **-0.0130 (0.008)** | -0.0056 (0.255) | **1.327 (0.013)** | 0.699 (0.171) | 0.356 (0.116) | | **1.047 (0.028)** | 0.319 (0.691) |
| Infant gut microbiome | PC1 | 0.0007 (0.948) | | -0.015 (0.25) | | -0.009 (0.113) | | -0.012 (0.3) | |
| | PC2 | 0.0008 (0.547) | | **-0.059 (0.005)** | **-0.039 (0.016)** | **-0.0169 (0.007)** | -0.011 (0.093) | **-0.046 (<0.001)** | **-0.038 (0.010)** |
| | BF ratio | 0.0003 (0.320) | | 0.004 (0.884) | | -0.005 (0.658) | | -0.002 (0.927) | |
| | FA ratio | 0.0001 (0.531) | | 0.021 (0.346) | | 0.004 (0.660) | | 0.017 (0.376) | |
| Maternal | Years in Canada | 0.0002 (0.220) | | **0.041 (0.014)** | | 0.011 (0.128) | | 0.012 (0.423) | |
| Perinatal | GDM | -0.0030 (0.672) | | **-1.491 (0.057)** | **-1.404 (0.077)** | **-0.683 (0.037)** | **-0.651 (0.056)** | -1.084 (0.120) | |
| | Delivery Mode | -0.0016 (0.596) | | -0.153 (0.828) | | -0.177 (0.547) | | **-1.328 (0.036)** | -0.367 (0.236) |
| | Maternal Antibiotic Use in pregnancy | -0.0016 (0.712) | | -0.662 (0.163) | | **-0.390 (0.050)** | **-0.381 (0.046)** | -0.550 (0.189) | |
| | Maternal Vegetarian Status in pregnancy | -0.0030 (0.541) | | -0.887 (0.104) | | -0.287 (0.210) | | -0.465 (0.337) | |
| Infant | Birth weight | **0.0075 (0.064)** | | -0.616 (0.174) | | -0.269 (0.155) | | **-1.035 (0.009)** | **-0.866 (0.452)** |
| | Ethnicity | **-0.0068 (0.096)** | -0.0081 (0.075) | **-1.395 (0.002)** | | **-0.430 (0.023)** | | -0.458 (0.256) | |
| | Sex | -0.0024 (0.549) | | 0.383 (0.401) | | 0.034 (0.860) | | 0.371 (0.358) | |
| | weight gain in the 1st year | -0.0015 (0.305) | | 0.109 (0.506) | | 0.097 (0.156) | | **0.252 (0.082)** | 0.267 (0.076) |
| | Age | -0.0031 (0.033) | | 0.394 (0.015) | | **0.145 (0.045)** | 0.103 (0.132) | 0.372 (0.010) | |

| | | Lysine | | Methionine | | DMG |
|---|---|---|---|---|---|---|

| | Variable | Univar. | FSW | Univar. | FSW | Univar. | FSW |
|---|---|---|---|---|---|---|---|
| Infant diet | Breastfeeding Status at 1 year | **-0.127 (0.000)** | **-0.521 (0.021)** | **-0.127 (0.000)** | **-0.086 (0.013)** | **-0.093 (0.000)** | **-0.058 (0.005)** |
| | Time since Weaning | 0.039 (0.308) | | **0.005 (0.463)** | | 0.005 (0.255) | |
| | Formula Feeding Status at 1 year | -0.051 (0.813) | | -0.005 (0.906) | | **0.065 (0.003)** | **0.044 (0.037)** |
| | Age of Solid Food Introduction | 0.282 (0.583) | | -0.024 (0.805) | | 0.026 (0.631) | |
| | Cow Milk Consumption | **0.409 (0.075)** | 0.237 (0.281) | 0.046 (0.258) | | 0.021 (0.353) | |
| Infant gut microbiome | PC1 | **-0.016 (0.003)** | | **-0.002 (0.035)** | | -0.001 (0.332) | |
| | PC2 | **-0.026 (<0.001)** | **-0.021 (0.003)** | **-0.005 (< 0.001)** | **-0.003 (0.004)** | **-0.003 (0.001)** | **-0.002 (< 0.001)** |
| | BF ratio | -0.004 (0.738) | | 0.002 (0.439) | | **0.003 (0.032)** | |
| | FA ratio | 0.015 (0.136) | | **0.004 (0.016)** | | **0.002 (0.041)** | |
| Maternal | Years in Canada | **0.031 (< 0.001)** | | **0.004 (< 0.001)** | | **0.001 (0.055)** | |
| Perinatal | GDM | **-0.691 (0.040)** | | **-0.135 (0.026)** | -0.098 (0.085) | -0.045 (0.187) | |
| | Delivery Mode | -0.141 (0.641) | | -0.051 (0.347) | | -0.009 (0.764) | |
| | Maternal Antibiotic Use in pregnancy | -0.214 (0.304) | | -0.052 (0.166) | | 0.001 (0.984) | |
| | Maternal Vegetarian Status in pregnancy | **-0.726 (0.002)** | **-0.533 (0.022)** | **-0.124 (0.003)** | -0.070 (0.088) | -0.039 (0.103) | |
| Infant | Birth weight | -0.093 (0.635) | | -0.029 (0.414) | | -0.022 (0.269) | |
| | Ethnicity | **-1.051 (< 0.001)** | | **-0.160 (< 0.001)** | | **-0.056 (0.005)** | |
| | Sex | 0.050 (0.802) | | -0.0004 (0.989) | | 0.007 (0.715) | |
| | weight gain in the 1st year | -0.023 (0.75) | | 0.003 (0.814) | | 0.008 (0.246) | |
| | Age | **0.143 (0.045)** | | **0.023 (0.072)** | | 0.001 (0.933) | |
