## Supplemental Table 3 for "Effects of Diet on the Microbiome and Serum Metabolome of South Asian Infants at 1 Year"

**Table S3**. Maternal and Infant Participant Characteristics for serum metabolite analysis.

| **Maternal Characteristics:** | | |
|---|---|---|
| | START (n=72) | CHILD (n = 81) |
| Antibiotic usage during labour<br>Yes<br>No | <br>33(45.8%)<br>36(50.0%)[b] | <br>33(40.7%)<br>44(54.3%)[b] |
| Vegetarian Status<br>Yes<br>No | <br>31(43.1%)<br>40(55.6%)[b] | <br>3(3.7%)<br>78(96.3%) |
| GDM<br>Yes<br>No | <br>8(11.1%)<br>64(88.9%) | <br>6(7.4%)<br>75(92.6%) |
| Years in Canada (SD) | 5.7(4.2) | 29.44(8.4) |
| **Infant Characteristics:** | | |
| Age in months at time of sample collection (SD) | 12.14(1.27) | 12.15(1.50) |
| Participant Sex<br>Male<br>Female | <br>34(47.2%)<br>38(52.8%) | <br>49(60.5%)<br>32(39.5%) |
| Delivery Mode<br>Vaginal<br>C-Section (planned)<br>C-Section (emergency) | <br>49(68.1%)<br>9(12.5%)<br>13(18.1%)[b] | <br>63(77.8%)<br>9(11.1%)<br>6(7.4%)[b] |
| Gestational Age in Weeks (SD) | 39.23(1.42) | 39.58(1.33) |
| Birthweight (Kg) | 3.29(0.49) | 3.55(0.49) |
| Weight gain (Kg) | 6.95(1.46) | 6.38(1.27) |
| Time in Months of introduction of solids:<br>0-3<br>3-6<br>6-9<br>9-12 | <br>1(1.4%)<br>60(83.3%)<br>10(13.9%)<br>1(1.4%) | <br>5(6.2%)<br>38(46.9%)<br>27(33.3%)<br>1(1.2%)[d] |
| Time in months Since Weaning (SD) | 8.03(3.31) | 5.36(3.32) |
| Ever Breastfed During First Year<br>Yes<br>No | <br>69(95.8%)<br>3(4.2%) | <br>79(97.5%)<br>2(2.5%) |
| Currently Breastfed at Sample Collection Time<br>Yes<br>No | <br>31(43.1%)<br>41(56.9%) | <br>28(34.6%)<br>53(65.4%) |

| | | |
|---|---|---|
| Ever Formula Fed During First Year<br>Yes<br>No | <br>63(87.5%)<br>5(6.9%)[c] | <br>42(51.9%)<br>30(37.0%)[d] |
| Currently Formula Fed at Sample Collection Time<br>Yes<br>No | <br>27(37.5%)<br>45(62.5%) | <br>18(22.2%)<br>54(66.7%)[d] |
| Current Cow's Milk Consumption at Sample Collection Time<br>Yes<br>No<br>NA | <br>47(65.3%)<br>23(31.9%)<br>2(2.8%) | <br>30(37.0%)<br>23(28.4%)<br>28(34.6%) |

[a]where there is no superscript there was no missing data
[b]Less than 5.00% data missing
[c]Less than 10.00% data missing
[d]10–20% data missing
