## Supplemental Table 2 for "Effects of Diet on the Microbiome and Serum Metabolome of South Asian Infants at 1 Year"

**Table S2.** START Diet-Serum Metabolite Associations Across Multiple Tests

| Diet Pattern | VIP Identified Metabolites | ANOVA (p-value) | FDR Adjusted ANOVA |
|---|---|---|---|
| Formula and Dairy (n = 72) | Valine, DMG, Tryptophan betaine, Leucine, Tyrosine, Phenylalanine, Iso-Leucine, methyl-Cysteine, 2-hba, Methionine, Lysine, TMAO | Valine ($p < 0.001$), DMG ($p < 0.001$), Tryptophan betaine ($p < 0.001$), Leucine ($p < 0.001$), Tyrosine ($p < 0.001$), Phenylalanine ($p < 0.001$), Iso-Leucine ($p < 0.001$), methyl-Cysteine (0.002), 2-hba (0.002), Methionine (0.003), Lysine (0.01), TMAO (0.01) | **Valine ($p < 0.001$), DMG ($p < 0.001$), Tryptophan betaine (0.01), Leucine (0.002), Tyrosine(0.002), Phenylalanine (0.002), Iso-Leucine (0.004), methyl-Cysteine (0.007), 2-hba (0.007), Methionine (0.008), Lysine (0.011), TMAO (0.011)** |
| Animal Foods (n=41) | methyl-hist, oxo-proline, DMG, serine, glycine, choline | methyl-Histidine (0.06), oxo-proline (0.02), DMG (0.044), Serine (0.045), glycine (0.059), choline (0.063), | **methyl-Histidine (0.011), oxo-proline (0.031),** DMG (0.062), Serine (0.062) Glycine (0.077) Choline (0.08) |
| Plant Foods (n = 41) | Allo leucine, Hippuric acid, cysteinylglycine disulfide, Tryptophan betaine, Serine | Allo leucine (0.023), Hippuric acid (0.03) cysteinylglycine disulfide (0.088) tryptophan betaine (0.13) serine (0.2) | **Allo leucine (0.037), Hippuric acid (0.046),** cysteinylglycine disulfide (0.10), tryptophan betaine (0.14), serine (0.21) |
